# Development of a Predictive Score for COVID-19 Diagnosis based on Demographics and Symptoms in Patients Attended at a Dedicated Screening Unit

**DOI:** 10.1101/2020.05.14.20101931

**Authors:** Alexandre P. Zavascki, Marcelo B. Gazzana, João Pedro M. Bidardt, Patrícia S. Fernandes, Aline Galiotto, Cristiane T. Kawski, Luiz Antonio Nasi, Diego R. Falci

## Abstract

**Background:** The diagnosis of COVID-19 based on clinical evaluation is difficult because symptoms often overlap with other respiratory diseases. A clinical score predictive of COVID-19 based on readily assessed variables may be useful in settings with restricted or no access to molecular diagnostic tests.

**Methods:** A score based on demographics and symptoms was developed in a cross-sectional study including patients attended in a dedicated COVID-19 screening unit. A backward stepwise logistic regression model was constructed and values for each variable were assigned according to their β coefficient values in the final model. Receiver operating characteristic (ROC) curve was constructed and its area under the curve (AUC) was calculated.

**Results:** A total of 464 patients were included: 98 (21.1%) COVID-19 and 366 (78.9%) non-COVID-19 patients. The score included variables independently associated with COVID-19 in the final model: age ≥60 years (2 points), fever (2), dyspnea (1), fatigue (1 point) and coryza (-1). Score values were significantly higher in COVID-19 than non-COVID-19 patients: median (Interquartile Range), 3 (2–4), and 1 (0–2), respectively; P<0.001. The score had an AUC of 0.80 (95% Confidence Interval [CI], 0.76–0.86). The specificity of scores ≥4 and ≥5 points were 90.4 (95%CI, 87.0–93.3) and 96.2 (95%CI, 93.7–97.9), respectively.

**Conclusions:** This preliminary score based on patients’ symptoms is a feasible tool that may be useful in setting with restricted or no access to molecular tests in a pandemic period, owing to the high specificity. Further studies are required to validate the score in other populations.

## INTRODUCTION

The coronavirus disease 2019 (COVID-19) epidemic has caused major public health impact in several countries [1, 2]. There has been a rapid progress in the characterization of clinical features of COVID-19; however, so far, most studies have focused on the evaluation of confirmed cases, without exploration of clinical findings that may distinguish COVID-19 from other respiratory conditions [3–5].

Recently, the Centers for Disease Control and Prevention (CDC) has added new symptoms(myalgias, chills, sore throat, new loss of taste or smell and headache), to its existing list of symptoms for COVID-19 (fever, cough, shortness of breath or difficulty breathing) [6]. However, although they may be frequent in COVID-19, as other symptoms previously in the list, these symptoms are common in other viral and even non-viral respiratory diseases [7]. Thus, any of the aforementioned COVID-19 symptoms individually seems to be specific for the disease. On the other hand, some symptoms that are common to other viral respiratory illnesses have not been frequently reported by COVID-19 patients, such as sneezing and coryza [3, 4]. The objective evaluation of the entire plethora of symptoms presented by the patients by a score may help to distinguish COVID-19 from other febrile or respiratory diseases.

A clinical score predictive of COVID-19 based on age, gender and symptoms may be particularly useful in, but not limited to, settings with restricted or no access to molecular diagnostic tests. A COVID-19 score based only on symptoms would be an easy, low cost and point of care diagnostic tool with many potential benefits to assist decision-process making of both primary-care and emergency department physicians. This point-of-care clinical diagnostic tool has been considered a research priority and has not yet been developed [8, 9]. Moreover, it may subsidize decisions on hospital infection control measures, home isolation and return to professional activities and contribute to diagnostic accuracy in telemedicine consults. Finally, it may contribute for epidemiological surveillance in low-income by increasing the specificity of the classification of “probable cases”.

In this study, we develop a score predictive for COVID-19 diagnosis based on demographic data (age and gender) and symptoms in patients attended for suspected COVID-19 in a dedicated screening unit.

## METHODS

### Study design and definitions

This study follows the Transparent Reporting of a multivariable prediction model for Individual Prognosis Or Diagnosis (TRIPOD) statement recommendations [10]. A retrospective cross-sectional study was performed at Hospital Moinhos de Vento, a private tertiary-care hospital from Porto Alegre, a 1.4 million inhabitants Brazilian city. All patients with suspected COVID-19 attended from January 28th to April 13rd at HMV were eligible for the the study.

From January 28th to March 14th, 2020, patients were considered suspected of COVID-19 if they presented fever or any respiratory symptom and have returned from countries with confirmed COVID-19 cases in the last 14 days. After March 14th, since community transmission has been documented in Brazil, travel history was not necessary for considering suspected of COVID-19. However, owing to restricted molecular tests availability, only patients with fever or respiratory symptoms with a more severe presentation (dyspnea, tachypnea, or low oxygen saturation, hypotension or any complementary laboratorial or radiological exams that might indicate severity) were tested for COVID-19.

Suspected patients were attended by a dedicated medical and nursery team in a specific COVID-19 screening area of the emergency department. Symptoms were collected from medical records (done for assistance purposes) of the first attendance.

The variables investigated were age, gender, fever, cough, sore throat, dyspnea, coryza, nasal congestion, sneezing, fatigue, myalgia, headache, diarrhea and nausea. A symptom was considered present if it were clearly registered either by spontaneous report of the patient or after specific questioning by the physician. Symptoms were considered absent either if there was a clear register of its absence or it has not been registered at all. Researchers registering the symptoms in the database were blinded to the outcome (COVID-19 status) since data collection from medical records were done in the same day or the day after the consultation before RT-PCR results were available.

The study included consecutive patients attended during COVID-19 pandemic and no sample size was previously defined. There was no missing data, since when not recorded, a symptom was considered absent. The study was approved by the institutional ethical committee (protocol number 4.018.709)

### Molecular diagnosis of SARS-CoV-2

Confirmation of COVID-19 diagnosis was done with one positive reverse-transcriptase polymerase chain reaction (RT-PCR) performed as previously reported [11]. RT-PCR. Patients with one or more negative RT-PCR were assigned to the non-COVID-19 group.

### Statistical analysis

Statistical analysis was performed using PASW Statistics for Windows v.18.0 (SPSS Inc., Chicago, IL). Bivariate analyses were performed using χ2 test or Fisher’s exact test for categorical variables and Student’s t-test for age. Age was further categorized based on its distribution in COVID-19 patients. Prevalence ratio (PR) and 95% Confidence Interval (CI) was calculated for each variable.

A backward stepwise logistic regression model was constructed with variables with a P ≤0.05 in bivariate analysis. Variables with a P > 0.05 were removed from the model at each step according to their P values (the variable with the highest P value at each step). Variables with a P ≤0.05 remained in the final model. Specific variables of clinical interest were allowed to be forced one by one into the final model, if they have not remained statistically significant in the backward stepwise model. They were planned to keep in the final model if it affects the β coefficient of any of the variables by ≥10%. Goodness of fit was assessed by Hosmer-Lemeshow test.

The value of each variable in score were assigned by rounding up β coefficient values of the final model to the first whole number. We also evaluated the performance of the score by assigning the exact value of β coefficient to each variable. Receiver operating characteristic(ROC) curve was constructed and its area under the curve (AUC) was calculated.

A preliminary validation was performed in a randomly selected sample with 25% of patients of the score development population. ROC curves and AUC were also calculated for this sample.

## RESULTS

In the development phase, 927 patients were attended at the dedicated emergency unit and collected a RT-PCR. Two-hundred nighty-three (31.6%) were excluded because they were health-care professionals, 68 (7.3%) were under 18 years-old and 102 (11.0%) were asymptomatic. A total of 464 patients were included in the study: 98 (21.1%) and 366 (78.9%) in COVID-19 and non-COVID-19 groups, respectively. Thirty-five (9.6%) patients from non-COVID-19 group had two RT-PCR tests from different days yielding negative results. Most patients (53%) were female and mean age was 48 years-old, but COVID-19 was significantly more frequent in male and mean age was significantly higher in these patients (Table 1). The most common symptom was cough, which was present in 67.3% of the patients, followed by fever (51.3%) and sore throat (36.2%). Headache, myalgias, dyspnea and coryza were present in 21 to 28% of patients, while fatigue (15.5%), nasal congestion (8.2%), diarrhea (7.3%) and nausea (5.8%) were the less frequent reported symptoms (table 1). Only three patients reported sneezing and it was not considered in bivariate analysis. Fever, dyspnea and fatigue were significantly more frequent in COVID-19 patients than non-COVID-19, while sore throat, headache, coryza and nasal congestion were significantly more common among non-COVID-19 patients (Table 1). The was no significant difference between groups in the frequency of cough, diarrhea and nausea.

**Table 1.**
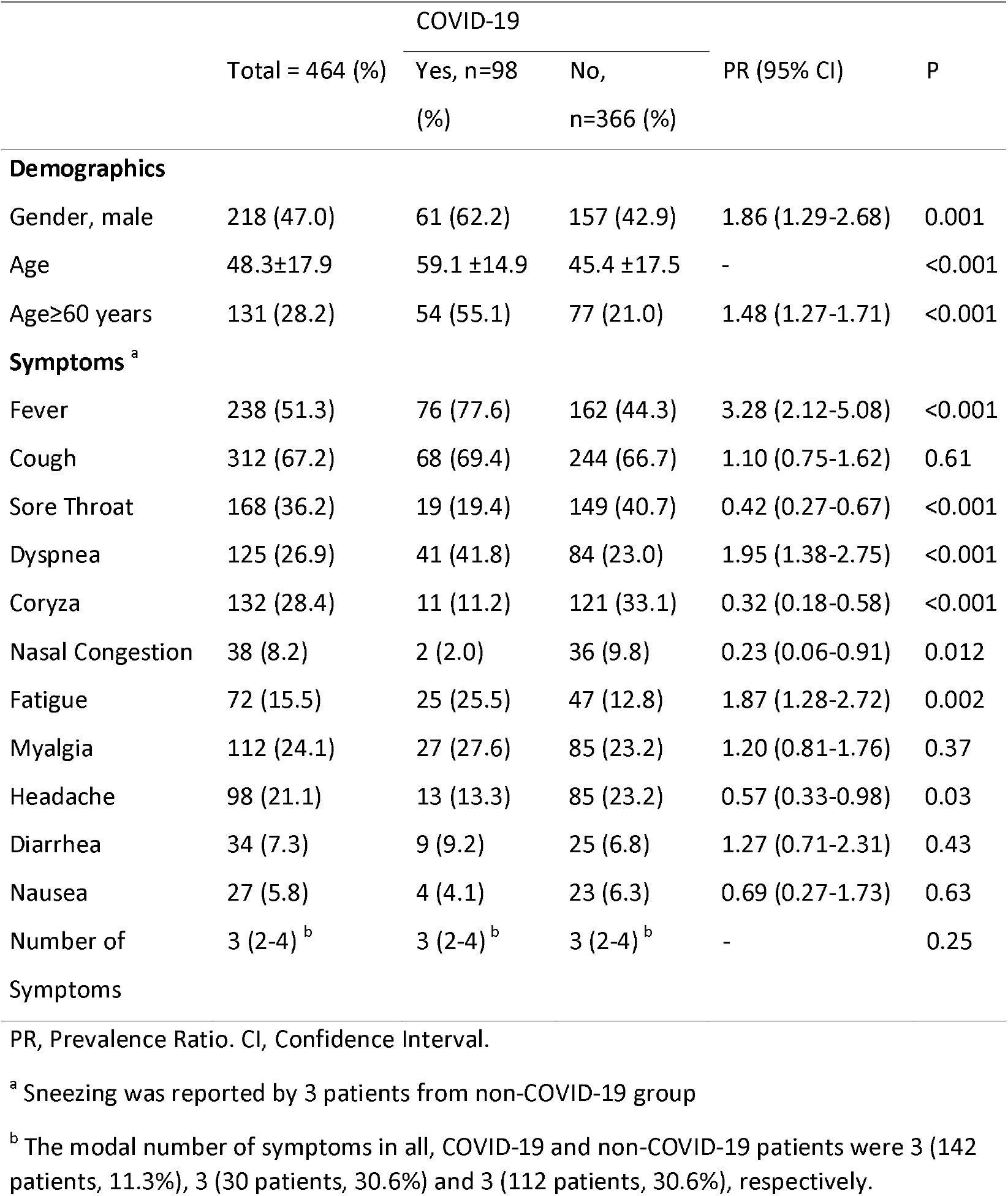
Bivariate analysis of demographics and symptoms in COVID-19 and non-COVID-19 patients.

|  | Total = 464 (%) | COVID-19 |  | PR (95% CI) | P |
| --- | --- | --- | --- | --- | --- |
|  |  | Yes, n=98 (%) | No, n=366 (%) |  |  |
| Demographics |  |  |  |  |  |
| Gender, male | 218 (47.0) | 61 (62.2) | 157 (42.9) | 1.86 (1.29-2.68) | 0.001 |
| Age | 48.3±17.9 | 59.1 ±14.9 | 45.4 ±17.5 | - | <0.001 |
| Age≥60 years | 131 (28.2) | 54 (55.1) | 77 (21.0) | 1.48 (1.27-1.71) | <0.001 |
| Symptoms <sup>a</sup> |  |  |  |  |  |
| Fever | 238 (51.3) | 76 (77.6) | 162 (44.3) | 3.28 (2.12-5.08) | <0.001 |
| Cough | 312 (67.2) | 68 (69.4) | 244 (66.7) | 1.10 (0.75-1.62) | 0.61 |
| Sore Throat | 168 (36.2) | 19 (19.4) | 149 (40.7) | 0.42 (0.27-0.67) | <0.001 |
| Dyspnea | 125 (26.9) | 41 (41.8) | 84 (23.0) | 1.95 (1.38-2.75) | <0.001 |
| Coryza | 132 (28.4) | 11 (11.2) | 121 (33.1) | 0.32 (0.18-0.58) | <0.001 |
| Nasal Congestion | 38 (8.2) | 2 (2.0) | 36 (9.8) | 0.23 (0.06-0.91) | 0.012 |
| Fatigue | 72 (15.5) | 25 (25.5) | 47 (12.8) | 1.87 (1.28-2.72) | 0.002 |
| Myalgia | 112 (24.1) | 27 (27.6) | 85 (23.2) | 1.20 (0.81-1.76) | 0.37 |
| Headache | 98 (21.1) | 13 (13.3) | 85 (23.2) | 0.57 (0.33-0.98) | 0.03 |
| Diarrhea | 34 (7.3) | 9 (9.2) | 25 (6.8) | 1.27 (0.71-2.31) | 0.43 |
| Nausea | 27 (5.8) | 4 (4.1) | 23 (6.3) | 0.69 (0.27-1.73) | 0.63 |
| Number of Symptoms | 3 (2-4) <sup>b</sup> | 3 (2-4) <sup>b</sup> | 3 (2-4) <sup>b</sup> | - | 0.25 |

PR, Prevalence Ratio. CI, Confidence Interval.
<sup>a</sup> Sneezing was reported by 3 patients from non-COVID-19 group
<sup>b</sup> The modal number of symptoms in all, COVID-19 and non-COVID-19 patients were 3 (142 patients, 11.3%), 3 (30 patients, 30.6%) and 3 (112 patients, 30.6%), respectively.

In the backward stepwise logistic regression model, the first variable excluded was headache and the last was gender (Supplemental Table 1). The variables that remained in the final model and their respective β coefficient and values assigned in the score are presented in table 2.

**Table 2.** Final logistic regression model and point values assign to each variable in the score.

| Variables <sup>a</sup> | $\beta$ coefficient | OR (95% CI) | P | Points assigned in the score | |
| --- | --- | --- | --- | --- | --- |
|  |  |  |  | Rounding up | Exact |
| Age≥60 years | 1.461 | 4.31 (2.58-7.20) | <0.001 | 2 | 1.4 |
| Fever | 1.397 | 4.04 (2.31-7.09) | <0.001 | 2 | 1.3 |
| Dyspnea | 0.580 | 1.79 (1.05-3.05) | 0.03 | 1 | 0.5 |
| Coryza | -1.000 | 0.37 (0.18-0.75) | 0.01 | -1 | -1 |
| Fatigue | 0.73 | 2.08 (1.12-3.85) | 0.03 | 1 | 0.7 |

OR, Odds Ratio. CI, Confidence Interval.
<sup>a</sup> Cough, myalgia, headache and sore throat were forced individually into the final model, but none has changed at least 10% the $\beta$ coefficient value.

The median (interquartile range [IQR]) and modal score values among all patients were 2 (0–3) and 2, respectively. The score distribution was significantly higher in COVID-19 than non-COVID-19 patients: median (IQR) and mode, 3 (2–4) and 3; and 1 (0–2) and 2, respectively; P<0.001 (Supplement Table 2). There was no significant difference in the AUC generated by ROC curves of rounded up and exact scores: 0.80 (95% CI, 0.76–0.86) and 0.81 (95% CI, 0.77–0.86), respectively.

Sensitivity, specificity, positive and negative likelihood ratio are displayed in table 3. Positive and negative predictive values for patients with score values ≥4 and ≥5 according to distinct COVID-19 prevalence are shown in Supplement Table 4.

**Table 3.** Specificity, Sensitivity, Positive and Negative Likelihood Ratios of COVID-19 score.

| Score Value <sup>a</sup> | Specificity (95% CI) | Sensitivity (95% CI) | PLR (95% CI) | NLR (95% CI) |
| --- | --- | --- | --- | --- |
| ≥0 | 14.5 (11.0-18.5) | 98.0 (92.8-99.8) | 1.15 (1.09-1.21) | 0.14 (0.03-0.57) |
| ≥1 | 36.3 (31.4-41.5) | 96.9 (91.2-99.4) | 1.52 (1.40-1.66) | 0.08 (0.03-0.26) |
| ≥ 2 | 53.0 (47.8-58.2) | 91.8 (84.6-96.4) | 1.95 (1.73-2.21) | 0.15 (0.08-0.30) |
| ≥ 3 | 76.8 (72.8-81.0) | 70.4 (60.3-79.2) | 3.03 (2.42-3.80) | 0.39 (0.28-0.53) |
| ≥ 4 | 90.4 (87.0-93.3) | 41.8 (32.0-52.2) | 4.37 (2.96-6.48) | 0.64 (0.54-0.76) |
| ≥ 5 | 96.2 (93.7-97.9) | 17.3 (10.4-26.3) | 4.53 (2.32-8.87) | 0.86 (0.78-0.94) |
| 6 | 99.5 (98.0-99.9) | 5.1 (1.7-11.5) | 9.34 (1.84-47.40) | 0.95 (0.91-1.00) |

PLR, Positive Likelihood Ratio. NLR, Negative Likelihood Ratio. CI, Confidence Interval.
<sup>a</sup> A score value of -1 has a specificity of 98.0% (95% CI, 92.8-99.8) for non-COVID-19 diagnosis. Sensitivity was 14.5% (95% CI, 11.0-18.5). RLR for non-COVID-19 diagnosis: 7,10 (95% CI, 1.76-28.6) and NLR: 0.87 (95% CI, 0.83-0.92)

For a preliminary validation, a sample of 124 patients were selected: 24 (19.4%) and 100 (80.6%) in COVID-19 and non-COVID-19 groups, respectively. AUC generated by ROC curves of score was 0.88 (95% CI, 0.81–0.96).

## DISCUSSION

Our study showed that a score based on symptoms readily recovered from the medical interview showed a high specificity to predict COVID-19. Specificity for COVID-19 was high in patients with scores ≥4 (90%) with some increase in scores ≥5 (96%) and 6 (99%), but with substantial decrease in the sensitivity for these latter values. PPV and NPV estimated for distinct COVID-19 prevalence scenarios (Supplemental Figure 1) showed that patients with either score values ≥4 and ≥5 may be considered as having probable diagnosis of COVID-19 when the prevalence among patients attended for fever or respiratory symptoms is high(>70%), and those with scores <4 may be provisionally excluded from having COVID-19 when a low prevalence (<10%) of COVID-19 is expected.

The comparative analysis between COVID-19 and non-COVID-19 patients is of paramount importance to define a clinical picture that may help physicians in the diagnosis. Notably, we showed that cough, the most common symptoms reported in several series [3, 4, 12], was also common among non-COVID-19 patients. Interestingly, other symptoms recently included in the CDC list of COVID-19 symptoms, such as headache and sore throat, were, in fact, more frequent in non-COVID-19 patients. Thus, although these symptoms may be present in COVID-19, they seemed to be not useful for the diagnosis in our study. On the other hand, fatigue, which is not specifically listed by CDC, was independently associated with COVID-19 in our study.

A previous study also evaluated predictive models of COVID-19, using epidemiological and clinical risk factors from a cohort of 54 COVID-19 cases and 734 controls from Singapore [13]. Of note, the authors have developed four prediction models including different combinations of clinical findings, demographic variables, laboratory tests (complete blood count, renal function tests and electrolytes) and radiologic findings (chest X-ray and/or chest computed tomography scan). The models had different performances according to the variables included. However, in model 4 (with only clinical information and demographic variables), predictive accuracy was reduced substantially (AUC 0.65; 95% CI, 0.57–0.73) [13]. The study presented here had important differences from this published work. Since our main objective was to subside rapid decisions in settings with restricted access to tests, such as primary care and emergency room triage (including telemedicine assessments), we did not include any laboratory or radiologic tests. Even though, our model, based only in demographic data and symptoms, performed very well as a discriminatory tool for suspected COVID-19 patients, and a diagnostic score was proposed based on model results.

Our study has several limitations that must be underlined. First, it was based on retrospective data from medical records and although clinical interview has been made by dedicated physicians, there was no standard questionnaire for it. So, it is possible that a less perceived symptoms that has been not specifically questioned by the physician might be missed. On the other hand, it is less likely that the main complains have been forgotten by the patients and not spontaneously referred. Second, symptoms not registered in medical records were assumed as not existent, when in fact there might be just not recorded by the physician. Third, we excluded COVID-19 based on only one negative test in most patients, which turns possible that in non-COVID-19 group there may be missed patients who would become positive with a second test. Nonetheless, this potential bias would decrease the specificity of the score. We have also not evaluated routinely and standardly other infectious etiology for patients’ diseases thus we were limited to classified patients as non-COVID-19. However, it does not affect our results considering the study purpose. Fifth, some symptoms recently added to the COVID-19 list have not been investigated such as chills, hyposmia and dysgeusia. We believe that especially the latter two symptoms have the potential to increase the score specificity and must be further evaluated [14]. Finally, performance of the score was satisfactory in the preliminary validation carried out in this study; nonetheless, it was not done in an external population and further validation is required. It was also conducted in a non-flu season (early autumn), and the score needs to be validated when influenza is more common.

In conclusion, we developed an early COVID-19 score based on patients’ symptoms with high specificity for the disease among patients attended at dedicated COVID-19 screening unit for fever or respiratory symptoms. We emphasize that the score may not be assumed as definitive. However, this preliminary score may be useful in setting with restricted or no access to molecular tests in a pandemic period, owing to the high specificity. Further studies are required to validate the score in other populations and in a flu season period and improve it with other clinical findings not assessed in this study.

## Data Availability

We hereby choose to not share our dataset.

## Acknowledgments

The authors thank Dione M. de Souza, Lindayane D. Motta, Lisiane R. Martins, Paulo F. B. Teixeira and Patricia M. Gleit for their hard work and valuable contribution in data collection, and Dr. Gisele Nader for her assistance in local regulatory issues. A. P. Z. is a research fellow of the National Council for Scientific and Technological Development (CNPq), Ministry of Science and Technology, Brazil.

